# A unique gene expression signature in visceral adipose tissue identifies a high blood pressure group in patients with Cushing’s syndrome

**DOI:** 10.1101/2023.02.28.23286599

**Authors:** Ulrich Stifel, Frederick Vogel, Giorgio Caratti, Martin Reincke, Jan Tuckermann

**Affiliations:** Institute of Comparative Molecular Endocrinology (CME), Ulm University, Ulm, Germany; Medizinische Klinik und Poliklinik IV, University Hospital, LMU Munich, Munich, Germany; Oxford Centre for Diabetes, Endocrinology and Metabolism, University of Oxford, UK

**Keywords:** Cushing’s syndrome, adipose tissue, hypertension, renin, cortisol, ACTH, adrenal hyperfunction, Cushing’s disease

## Abstract

Cushing’s syndrome (CS) is a rare disease caused by excess cortisol levels with high cardiovascular morbidity and mortality. Hypertension is a frequent feature of Cushing’s syndrome, promoting hypercortisolism-associated cardiovascular events. Adipose tissue is a highly plastic tissue with most of the major cell types strongly affected in their function by the excess cortisol exposure. We hypothesized that the molecular and cellular changes of visceral adipose tissue (VAT) in response to cortisol excess can impact on systemic blood pressure levels in patients with CS. We, therefore, investigated gene expression signatures in VAT from patients with CS collected during curative adrenal surgery identifying significant alterations. During active CS we observed a strong downregulation of gene programs associated with immunity and inflammation in the VAT. In addition, we observed an clustering of the patients based on VAT gene expression profiles into two groups (CS^Low^ and CS^High^) according to blood pressure levels. The two clusters showed significant differences in gene expression pattens of the renin-angiotensin-aldosterone-system (RAAS). Renin (REN) was the strongest regulated gene compared to control patients and its expression correlated with increased blood pressure observed in our patients while systemic renin plasma levels were suppressed indicative of an abnormal blood pressure and volume status in response to VAT RAAS activation. Here we show for the first time a relevant contribution of the local RAAS system on systemic blood pressure levels in patients with CS. Patients from the CS^High^ group had still a significant increased blood pressure levels 6 months into remission, highlighting the importance of local tissue effects on long-term systemic effects observed in CS.

## Introduction

Endogenous Cushing’s syndrome (CS) is a rare disease affecting 1-3 people per million per year [1]. It is caused by chronic excessive secretion of cortisol by the adrenal glands. Consequently, patients have increase morbidity and mortality, and this is mainly due to cardiovascular and cerebrovascular events as well as infectious complications. According to the European Cushing’s Registry ERCUSYN, mortality is highest around the time of diagnosis until 90 days following start of therapy [2]. In Denmark, patients with active CS had a seven-fold increased risk of venous thromboembolism because of hypercoagulable state, a six-fold increased risk of heart failure, and a four-to-five-fold risk of stroke, compared with the background population [3]. A nation-wide cohort study from Sweden demonstrated similar results, with increased cardiovascular morbidity starting up to 3 years prior diagnosis, and elevated risks for stroke and myocardial infarction even years after biochemical remission [4].

Up to 85% of patients with CS have hypertension at diagnosis, and 9% may require hospital admission because of hypertensive crisis [5]. Prevalence of hypertension depends on subtype, duration of hypercortisolism, age, and co-existing metabolic comorbidities, such as diabetes mellitus, obesity and other components of the metabolic syndrome [6-8]. Whether a sex-specific difference exists in hypercortisolism-associated hypertension remains controversial [9, 10]. Interestingly, no correlation between blood pressure values and cortisol levels was observed [11-13]. However, hypertension has been reported to be an independent factor associated with increased mortality in CS [14, 15].

The pathophysiology of CS-related hypertension is complex and not well understood. The supposed pathogenetic mechanisms include a broad spectrum of contributing factors, such as mineralocorticoid receptor activation, vasoactive factors, increased vascular reactivity to vasoconstrictors as well as activation and/or modulation of the renin-angiotensin system (RAS) [5, 11, 13, 16]. Since phenotypically the visceral adipose tissue (VAT) is massively increased in CS, and visceral obesity has a major impact on the clinical phenotype during active CS as well as the persistence of several comorbidities including hypertension [17-21], we assumed that auto- and paracrine factors of the VAT may be relevant for systemic blood pressure regulation [22-24]. Therefore, we hypothesize that factors from VAT, especially components of the RAS, can modulate systemic blood pressure levels in patients with CS, independent of pharmacological intervention. To prove our hypothesis, we studied VAT from patients with CS obtained during curative adrenal surgery and investigated the association between expression of the local RAS in VAT using RNAseq and immunohistochemistry, and the hypertension phenotype at time of diagnosis as well as following curative surgery.

## Patients and methods

### Patients

A total of 17 patients with an unilateral adrenocortical tumor who underwent adrenalectomy at the Ludwig Maximilians University Munich were included in this study. Of these, 12 patients were diagnosed with overt adrenal Cushing’s syndrome due to cortisol-producing adrenal adenoma (CPA) and 5 with nonfunctioning adrenal tumor (NFAT) who served as controls. All patients with adrenal CS had typical signs and symptoms at the time of diagnosis. Diagnosis and subtype differentiation were based on the criteria recommended by current clinical practice guidelines [1, 25], following standard operational procedures as reported previously [18, 26]. NFAT was confirmed by at least one normal biochemical test for endogenous hypercortisolism and the absence of typical hypercortisolism-associated signs and symptoms. Exclusion criteria were age above 70 or below 18 years, mild autonomous cortisol secretion, CS due to adrenocortical carcinoma, bilateral adrenal masses, persistent disease after surgery and adrenostatic therapy prior to surgery. Because of the higher prevalence of CPAs in women [27, 28], and thus more tissue samples available, only female patients were included in the study. The number of cases available and tissue samples stored during the inclusion period determined the sample size of female patients with overt adrenal CS and NFAT. Periadrenal adipose tissue was separated from the adrenal gland immediately after receipt of the resectate and stored at -80°C. Because of the limited availability of paraffin-embedded periadrenal adipose tissue, we took tissues of additional patients for histological analyses difered (CPA n=10, NFAT n=5).

Patients with CS underwent clinical and biochemical assessment at the time of diagnosis before surgery and again 6 months following adrenalectomy. Successful surgery outcome was confirmed by clinical and biochemical disease remission including the presence of adrenal insufficiency postoperatively. During the adrenal insufficiency phase, all patients received standard glucocorticoid replacement therapy, consisting of 20-25 mg hydrocortisone per day.

The study was performed as part of the German Cushing’s registry. Patients were diagnosed and underwent adrenalectomy between 2012 and 2021 at the Munich center of the German Cushing’s registry. The German Cushing’s registry (NeoExNet, No. 152-10) was approved by the LMU ethics committee, and all patients gave written informed consent.

### Blood pressure measurements

Blood pressure measurements were taken during the visits at the time of diagnosis and 6 months after successful surgery. Blood pressure was measured in a sitting position, using an automatic oscillometric device, after 5 minutes of rest. The mean value of three consecutive measurements was used in the analysis.

### Measurement of blood and serum parameters

Venous blood samples were taken after overnight fast at the time of diagnosis. Hormonal laboratory analyses included serum cortisol, aldosterone and plasma renin concentration, cortisol after overnight 1 mg suppression, urinary free cortisol measurements and late-night salivary cortisol determinations. All lab tests were performed in the endocrine laboratory, Department of Medicine IV, and in the Clinical Laboratory Department, University Hospital Munich. Measurements of serum aldosterone and renin concentrations were made in one run at the end of the inclusion period, reducing analytical variability. Further biochemical parameters were performed in the Institute of Laboratory Medicine, University Hospital Munich, using standard methods.

### RNA-seq

Visceral adipose tissue (VAT)-biopsy from 14 patients with adrenal CS and 5 with NFAT who served as controls were obtained after adrenalectomy, respectively. VAT was stored at -80°C and total RNA was isolated following the TRIZOL RNA isolation protocol (Invitrogen). Quality of RNA was analyzed by capillary electrophoresis on an Agilent System Agilent 2100 was determined. RNA-seq was performed by Novogen (UK) on an Illumina Novaseq 600. Data quality control was performed on the basis of Pearson correlation and samples with a low correlation (<0.6) were removed (2 samples). The paired-end data was aligned to the human genome GRCh38 (hg38) using hisat2 [29]. Differential expression analysis between the control group and the CS patients, as well as between the two groups of CS patients were performed using DESeq2 [30]. Differentially expressed genes (DEG) were defined using a p.value of <0.05 and a log2foldchange of >1 as criteria. Volcano plots for the DEG were generated using R (3.6.3 and 4.04). Heatmaps were generated using MORPHEUS (Morpheus, https://software.broadinstitute.org/morpheus), and ComplexHeatmap [31]. Data on protein origin (secreted, membrane bound, intracellular) was obtained from The Human Protein Atlas [32]. Principal component analysis was performed using the pcaExplorer package [33]. Pathway analysis was performed using the DEG previously defined with ENRICHR [34]. Top 4 hits are shown sorted by p.value for the Elsevier pathway analysis and the gene ontology biological process analysis. Epigenetic Landscape In Silico deletion Analysis (LISA) was performed using the top most 500 DEG, sorted by p.value [35]. Gene set enrichment analysis (GSEA) was performed using the GSEA 4.2.3 version of the local application [36]. In silico cytometry was performed using the Cibersortx web application [37]. For this purpose, signature matrices were generated using all the available VAT human single cell data of lean patients from [38] to estimate cell type populations in our bulk RNA-seq approach. Data was processed using Galaxy Europe [39].

### Sample Data Availability

Sequencing data was uploaded to the GEO database and is available under GSE218965.

### Immunohistochemistry

Sections with VAT from CS patients and controls were rehydrated in xylene and an alcohol series. Subsequently antigen retrieval in citrate-based antigen retrieval solution (Sigma, C999) for 20 minutes at 100°C was applied. Samples were blocked using a ready-to-use blocking solution (Milipore, 20773) and incubated with antibody (1:200 Cd68 abcam KP1, 1:100 PDE4D Sigma-Aldrich (SAB4502128)) diluted in blocking solution. Endogenous peroxidases were quenched with 0.03% v/v H_2_O_2_ for 20 mins after which the samples were blocked using avidin/biotin blocking kit (Vector Laboratories SP-2001). Sections where incubated with anti-rabbit, or anti-mouse biotinylated secondary antibody 1:800 (BA-1000 or BA-9200 Vector Laboratories) for 2 hours at 4°C. After washing with PBS, samples were incubated with streptavidin-conjugated horseradish peroxidase (SA-5004 Vector laboratories 1:300 in PBS 0.1% Tween-20) for 1 hour at 4°C. Color detection on the samples was performed using 3,3’-820 diaminobenzidine (ImmPACT DAB SK-4100, Vector Laboratories) and nuclei with Hematoxylin solution (H3401 Vector Laboratories). Sections were then dehydrated and mounted using Eukitt (Langenbrinck 04-0001). Images were acquired using 10x magnification on an Olympus BX41 microscope.

### Blood vessel quantification

Blood vessel were counted on whole tissue slides from each patient and the number of blood vessels was normalized to the whole tissue area.

### Image Analysis

Pictures from IHC were analyzed using ImageJ Fiji software [40]. Quantification was performed as stained area/total area of the pictures and shown in %.

### Statistical Analysis

Statistics were performed using GraphPad 9 and R (3.6.3 and 4.0.4). Categorical data was analyzed using Fisher exact test and continuous data was analyzed using Mann-Whitney test or two-way ANOVA. Correlations were performed using GraphPad with simple linear regression model. P.value of below 0.05 was consider a significant effect.

## Results

### Visceral Adipose Tissue transcriptome is altered by active Cushing’s syndrome

To clarify specific gene signatures in VAT of patients with Cushing’s syndrome we collected VAT from 12 patients with CPA and 5 patients with NFAT. Clinical data are shown in **Table 1**. As expected, patients with CS had higher blood pressure values compared to controls and an elevated homeostatic model assessment for insulin resistance index (HOMA-IR). Body mass index (BMI) and serum potassium were comparable in both groups.

**Table 1:** Patient characteristics at the time of diagnosis. Statistical significance was measured using Mann-Witney U test for continuous data and Fishers exact test for categorical data. Data were considered significant at p < 0.05*, p < 0.01**, p < 0.001***, p < 0.0001****. Abbreviations: BMI, Body mass index; WHR, Waist-to-hip ratio; WC, waist circumference; HbA1c, Hemoglobin A1c; HOMA-IR, homeostatic model assessment for insulin resistance; BP, blood pressure; LNSC, late night salivary cortisol; UFC, urinary free cortisol, LDDST, low dose dexamethasone suppression test. Data is shown as SEM ± standard deviation.

|  | Non-functional adenoma<br>(n=5) | Cushing's syndrome (n=12) | Reference | p.value |
| --- | --- | --- | --- | --- |
| Age (years) | 51 ± 5.4 | 44.5 ± 2.6 |  | 0.2448 |
| Sex (female/male) | (5/0) | (12/0) |  | 1 |
| BMI (kg/m<sup>2</sup>) | 27.36 ± 2.7 | 28.05 ± 2.0 |  | 0.9593 |
| WC (cm) | - | 93.07 ± 18.4 |  | - |
| WHR | - | 0.8417 ± 0.03 | < 0.85 | - |
| Fasting glucose (mg/dl) | 106.4 ± 11.6 | 93.50 ± 2.7 | 60 - 99 | 0.3410 |
| HbA1c (%) | 5.500 ± 0.2 | 5.750 ± 0.1 |  | 0.3559 |
| HOMA-IR | - | 3.769 ± 0.9 (n=11) |  | - |
| Diast. BP (mmHG) | 83.80 ± 1.9 | 92.40 ± 2.5 |  | 0.0493* |
| Syst. BP (mmHG) | 126.80 ± 5.2 | 142 ± 3.6 |  | 0.0209* |
| Serum Potassium (mmol/L) | 4.420 ± 0.16 | 4.167 ± 0.09 | 3.5 - 5.1 | 0.2015 |
| LNSC (ng/ml) | 0.9 ± 0.3 (n=3) | 10.78 ± 4.2 | ≤ 1.5 | 0.0044** |
| UFC (µg/24h) | 45 ± 0 (n=1) | 434.3 ± 108.4 | ≤ 83.0 | - |
| ACTH basal (pg/ml) | 10.60 ± 1.9 | 2.917 ± 0.4 | 4 - 50 | 0.0002*** |
| LDDST (µg/dl) | 1.7 ± 0.8 (n=3) | 16.94 ± 2.3 | ≤ 1.8 | 0.0044** |
| Cortisol basal | 9.380 ± 1.7 | 17.59 ± 2.3 |  | 0.0255* |

**E**xpression profiling by RNAseq identified 1073 genes differentially regulated as shown by the volcano blot (**Figure 1A)** and the principal component analysis (PCA) **(Figure S1A**). Elsevier Pathway analysis of these differentially expressed genes (DEG) showed hypertension and colitis as the most regulated pathways, while gene ontology analysis revealed a differentially regulation of protein folding dependent on chaperons (**Figure 1B**). The top term in the Pathway enrichment was arterial hypertension, which includes a wide variety of genes of which the differentially expressed ones include *REN, VWF* and *VEGFA* (**Figure 1C**). To investigate the potential regulators of these DEG, we performed an epigenetic landscape in silico deletion analysis (LISA) [35]. Here we found that the most likely transcription factors regulating the DEGs were not the suspected cortisol activated transcription factors GR (*NR3C1*) or MR (*NR3C2*), but rather EGR3, TERC, CTCF, and the ESR1 (estrogen receptor), in line with previously published data sets (**Figure S1B**) [41].

**Figure 1:**
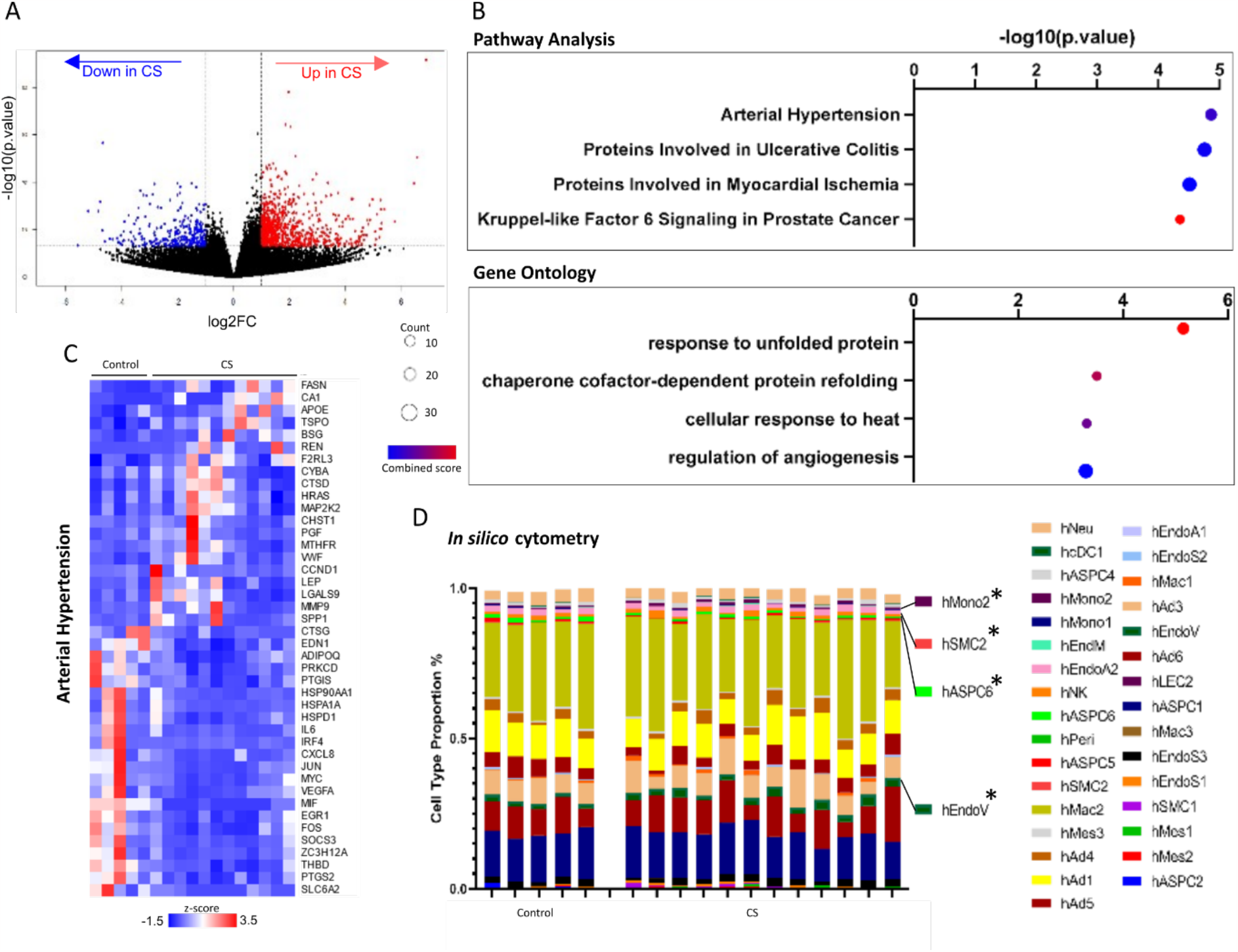
Transcriptome changes in female human patients during active adrenal Cushing syndrome disease in visceral adipose tissue and control patients with non-functional adenomas. A) Volcano plot from 1073 DEG between female CS patients and matched control patients p.value < 0.05, log2FC < 1. B) Elsevier Pathway analysis from DEG, top 4 terms shown and Gene Ontology biological process analysis from DEG, top 4 terms shown (response to unfolded protein (GO:0006986), chaperone cofactor-dependent protein refolding (GO:0051085), cellular response to heat (GO:0034605), regulation of angiogenesis (GO:0045765)). C) Genes of the top changed Pathway Analysis terms shown as heatmap with z-score D) In silico cell type distribution analysis using CibersortX from human patients from Emont *et al* 2022.

To further dissect the effects of active CS on adipose tissue we performed an *in silico* cytometry approach (Cibersortx), to estimate the putative changes in cell populations [37]. This approach revealed different abundance in four cell populations during active CS (**Figure 2A, C, E**). Monocytes are macrophage precursors from the blood stream and are actively infiltrating tissues, including the VAT [42]. Cibersortx analysis showed an increase of monocytes in the VAT from CS patients, this was further supported by IHC for CD68 and macrophage marker which was also increased in the tissue (**Figure 2B**). Strikingly gene set enrichment analysis (GSEA) from the controls versus the CS patients showed a decrease in TNFA signaling via NFKB in addition to many inflammation associated genes like *DUSP1, IL6* and *NFKB1* being downregulated and anti-inflammatory genes like *SPHK1* being upregulated.

**Figure 2:**
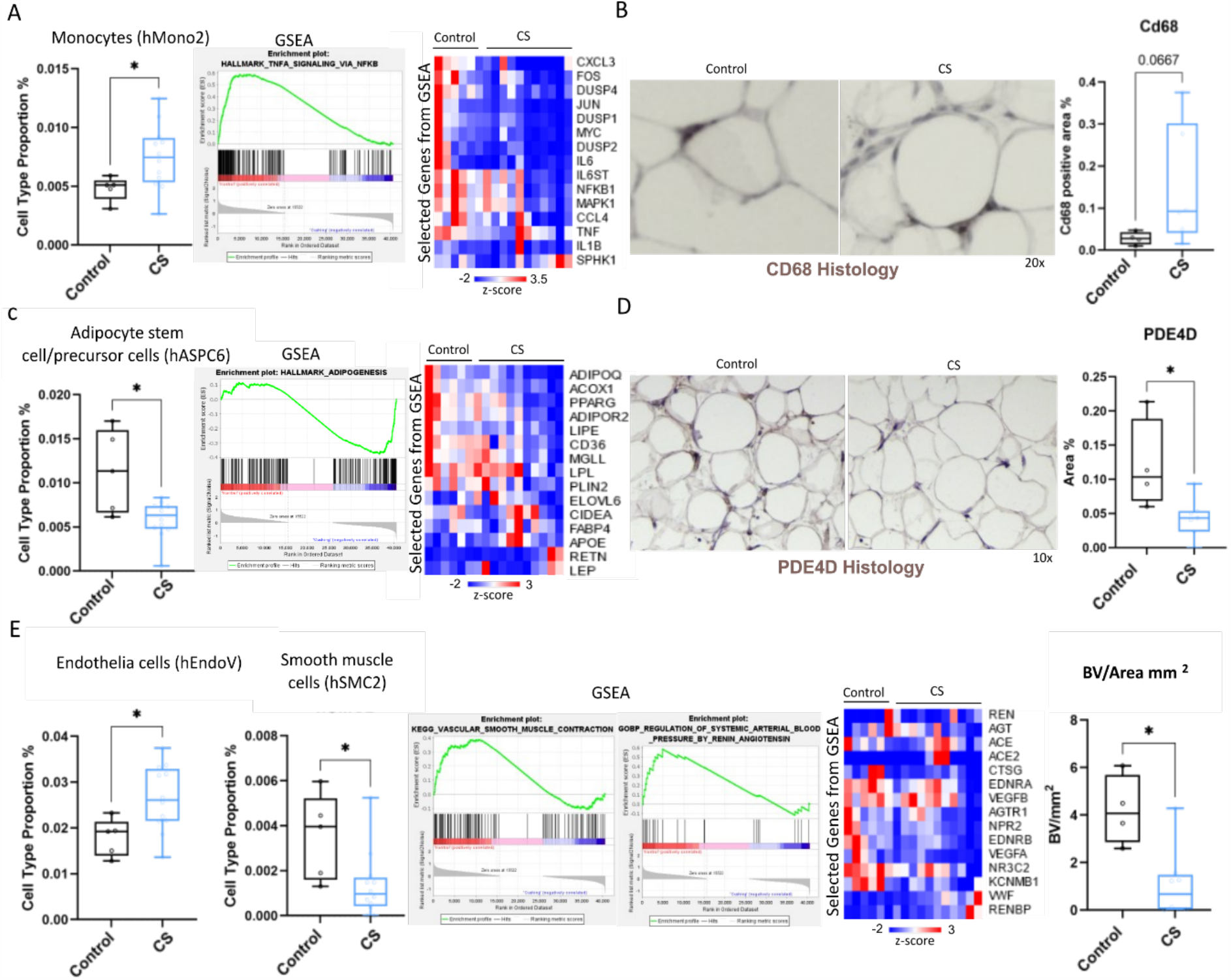
In silico analysis together with histology reveals differential cell populations in the VAT of female patients with CS and with non-functional adenomas as controls. A) Cell population of monocyte sup population 2, with GSEA between CS patients and control patients. Representative genes for the GSEA and cell population are shown. B) Histology for CD68 in patients with CS compared to control patients (n=4-6). C) Cell population of adipocyte stem cell and precursor population 6, with GSEA between CS and control patients. Representative genes for the GSEA and cell population are shown. D) Histology for PDE4D in patients with CS compared to control patients. E) Cell populations of human endothelia cells and human smooth muscle cells, with GSEA between CS and control patients. Representative genes for the GSEA and cell population are shown. Blood vessels / tissue area were counted. Statistical significance was measured using student t-test or Mann-Witney test. Data were considered significant at p < 0.05*, p < 0.01**, p < 0.001***, p < 0.0001****.

Furthermore, a fraction of the population of adipocyte stem- and precursor cells was downregulated in patients with active CS, which are defined as adipocyte stem- and precursor cells (hASPC6) (**Figure 2C**) [38], paralleled by downregulation in the GSEA for adipogenesis, including markers for adipocyte function and health, like *ADIPOQ, CD36* and *PPARG* (**Figure 2C**). This population of hASPCs was characterized by a high expression of *PDE4D*, a key marker gene for this cell population [38]. The expression of PDE4D was confirmed in the adipocyte stem- and precursor cell population from the CibersortX analysis (**Figure S2D**) and further by staining for it in the adipose tissue of the patients with IHC (**Figure 2D**).

Finally, we observed an increase in the abundance of endothelial cells and reduction of smooth muscle cells, suggesting an effect on the local vascular system. This is also confirmed by GSEA, which showed that the smooth muscle cell function and the local renin-angiotensin-aldosterone system (RAAS) were decreased (**Figure 2E**). In addition, we also found that the abundance of blood vessels (BV) was strongly reduced in the adipose tissue from patients with CS compared to NFAT control tissue (**Figure 2E**). This was also confirmed as many components of the RAAS like *REN, AGTR1* and *VWF* were regulated during CS. These data suggest a wide range of physiological consequences on the adipose tissue during CS, with major cell populations and important pathways and genes being affected.

### Gene expression reveals a correlation between VAT transcriptome and blood pressure

Unbiased clustering through PCA of the gene profiles of all samples indicated that the group of CS patients does not have a homogenous phenotype (**Figure S2A reordered from Figure S1A**). This is further confirmed by hierarchal clustering of the DEG between CS and NFAT control patients, in which we observed 3 clearly defined clusters (**Figure 3A**). Cluster 1 represented continuously downregulated genes and involved a high number of genes encoding secreted proteins, involved in resolution of inflammation and hypertension. In contrast mRNAs in cluster 2 and 3 were upregulated, but genes in cluster 3 are only upregulated in a CS subgroup. These subgroups of CS patients showed no difference in age, body composition, diabetic metabolic state or the severity of hypercortisolism measured by UFC, LNSC and LDDST (**Figure 3B, Figure S2B**). However, the subgroups of CS patients (Cluster 3 low and Cluster 3 high expression) were strongly different in their hypertensive phenotype during glucocorticoid excess. While the first subgroup (Cluster 3 low expressed, now designated as CS^Low^) was characterized by a non-significant, slightly higher blood pressure compared to the NFAT controls, the second subgroup (Cluster 3 high expressed, now designated as CS^High^) had significantly higher blood pressure levels with a pronounced hypertensive phenotype (**Figure 3B, Figure S2B**). This effect was independent of antihypertensive medication, as the number of patients receiving antihypertensive drugs was comparable in both subgroups of CS patients (CS^Low^: 4/6, CS^High^: 5/6) (**Figure S2C**).

**Figure 3:**
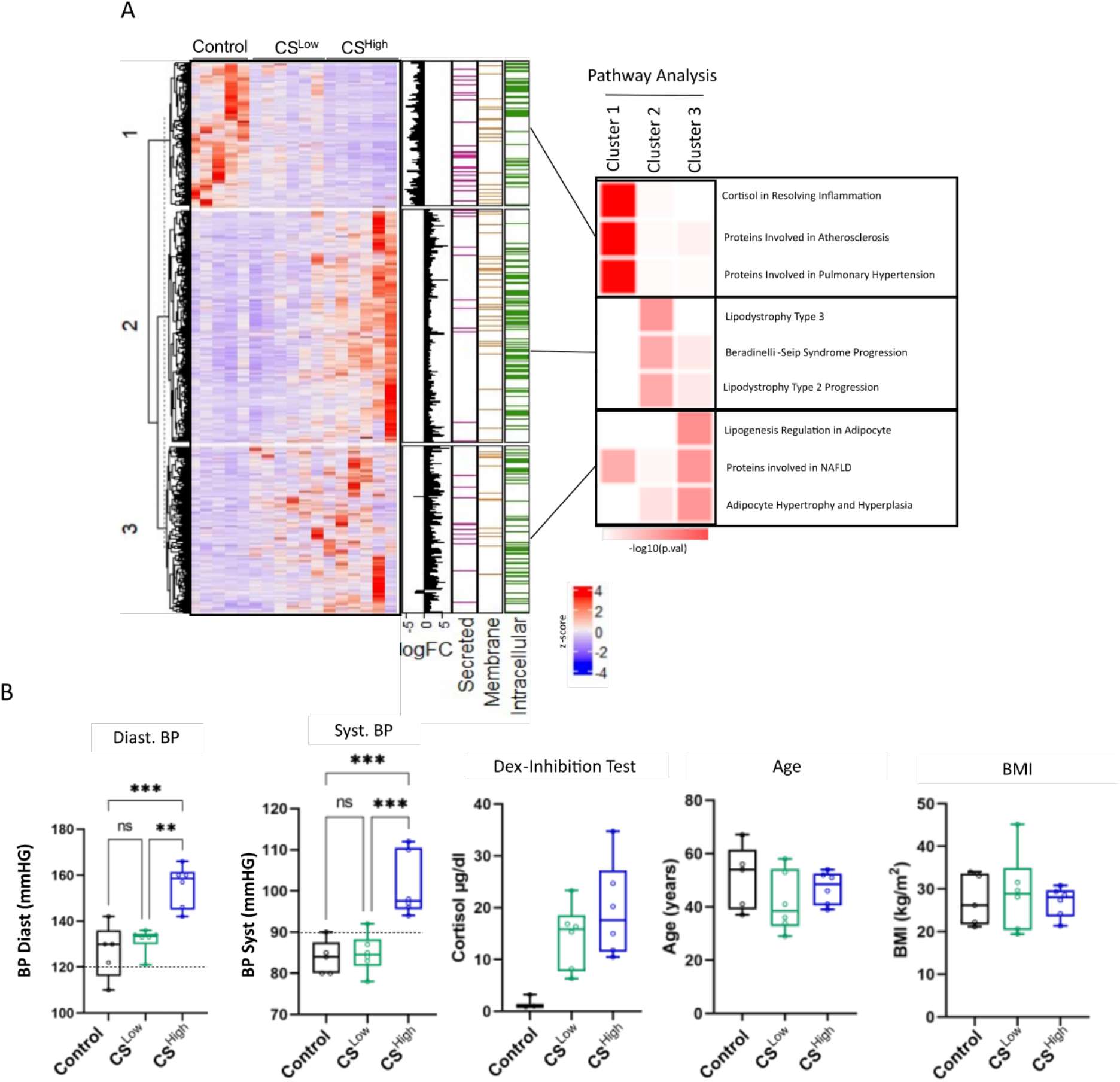
Visceral adipose tissue transcriptome of female CS patients reveals blood pressure heterogeneity. Female patients with non.functional adenomas served as controls. A) Heatmap showing the DEG between CS and control patients after hierarchical clustering with Pathway Analysis from the 3 identified clusters. B) Patient data shown as boxplots after separation into the 3 groups given by the clustering Statistical significance was measured using two-way ANOVA. Data were considered significant at p < 0.05*, p < 0.01**, p < 0.001***, p < 0.0001****.

Between the gene signatures assigned to the CS^Low^ group and CS^High^ group we observed 4883 DEGs. Pathway analysis revealed a high score of adiponectin synthesis and other adipokines (**Figure S3A, B**). Similar to the total comparison of controls with CS patients we found by LISA that neither GR (NR3C1) nor MR (NR3C2) were among the top candidates for potential TF regulating the DEG (**Figure S3C**). Rather PHF8, MAZ, CDK9 and MYC/MAX were among the relevant regulators.

The topmost regulated gene was renin (REN) responsible for the conversion of angiotensinogen to angiotensin I. The RAAS is known to be altered in CS [13]. This gene was also regulated according to the previously defined subgroups of patients with CS, although not reaching statistical significance (**Figure 4A**). Intriguingly the expression of RAAS components in VAT like renin and others was shown to be a major factor in obesity related blood pressure pathologies [24]. We found that the VAT gene expression of REN correlated significantly with the systolic blood pressure of all patients included in the study, with an R^2^ of 0.237 (**Figure 4B**). Plasma renin and aldosterone levels were not significantly different in CS patients compared with control patients, consistent with previous studies investigating the systemic RAAS in CS, which reported plasma renin and aldosterone concentrations within the normal or slightly below the normal range [13, 43-46]. Interestingly, plasma renin concentrations were significantly reduced in the CS^High^ versus the CS^Low^ subgroup, and the aldosterone to renin ratio elevated, indicating a systemic endocrine response of the renal juxtaglomerular apparatus to an abnormal high blood pressure and volume status (**Figure 4C left)**. Noteworthy, REN gene expression in VAT did not correlate with plasma renin concentrations measured in patients with CS (**Figure 4D**). Aldosterone, aldosterone to renin ratio and serum potassium levels were not different between subgroups and compared to controls (**Figure 4C right**). Moreover, plasma renin concentrations were not predictive of high blood pressure levels, as we observed no correlation between plasma renin levels and blood pressure (**Figure S3E**).

**Figure 4:**
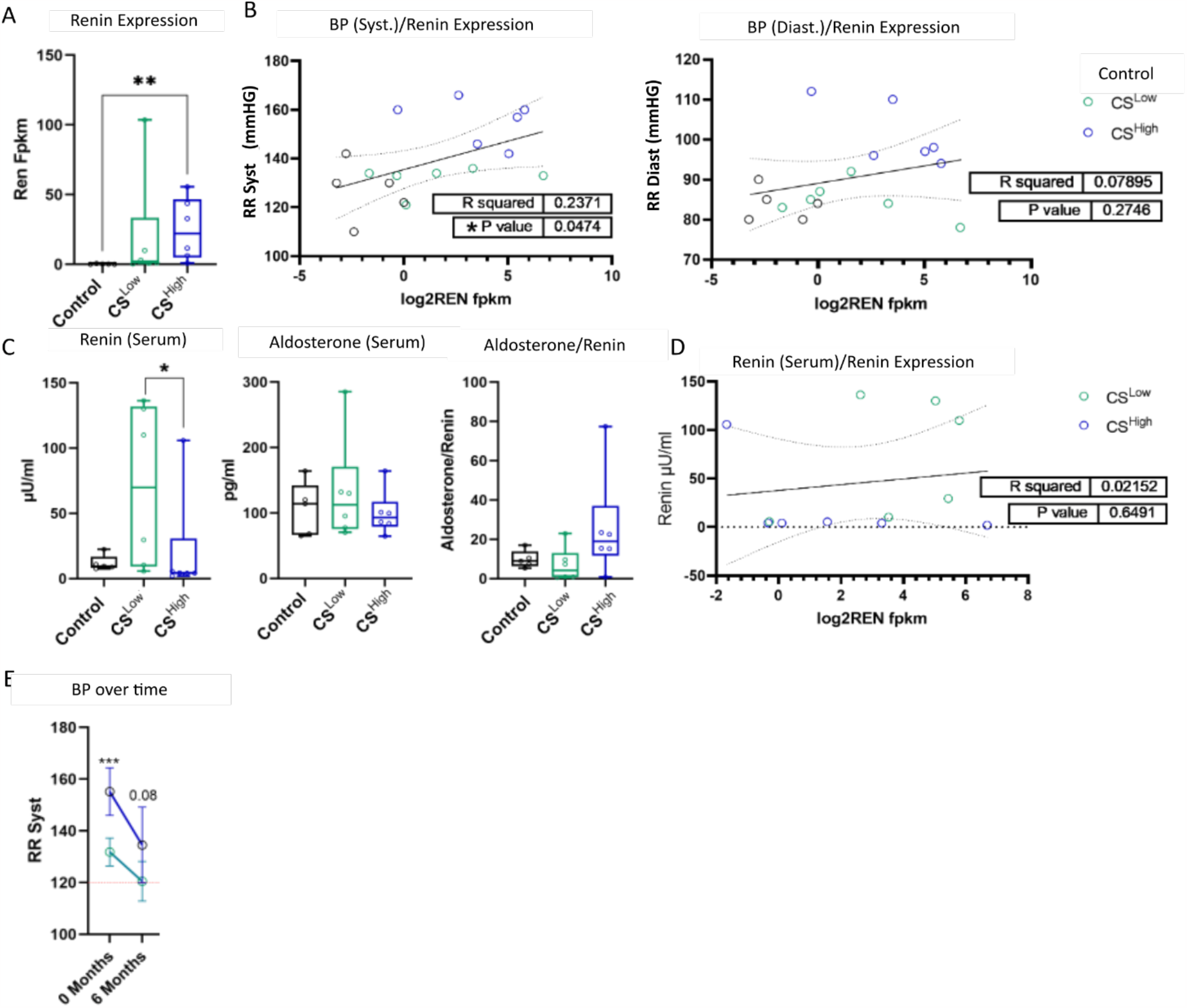
Renin and other components of the RAAS are regulated in VAT from a subset of female CS patients and control systemic blood pressure. **A**) Renin (REN) gene expression from Fpkm shown. B) Correlation between systemic systolic and diastolic blood pressure and REN (VAT) gene expression. C) Renin and aldosterone serum levels. D) Correlation between Renin VAT gene expression and Renin serum levels. E) Blood pressure decrease over time after surgery is shown for 6 months from both subgroups of CS patients. Statistical significance was measured using two-way ANOVA or Mann-Witney test. Data were considered significant at p < 0.05*, p < 0.01**, p < 0.001***, p < 0.0001****.

Six months following curative surgery, both subgroups showed lower blood pressure values compared with the time of diagnosis, as expected (**Figure 4F**). Remarkably, there was still a trend towards higher blood pressure values in patients of CS^High^ subgroup compared to CS^Low^ (p=0.08) after 6 months in remission (Patients on BP drugs CS^Low^: 1/6, CS^High^: 1/6). This indicates a potential long-lasting effects of adipose tissue RAS components despite sustained biochemical remission of CS.

## Discussion

Management of patients with CS and the thereof caused long-term complications are still a major challenge today [28, 47]. VAT taken as part of the adrenalectomy shows significant alterations in the transcriptome (**Figure 1**) compared to non-CS patients, as previously reported [41, 48]. Consistent with previous studies, our study highlights the profound effects of chronic cortisol excess on adipose tissue gene expression. We discovered a unique gene expression signature in VAT that is associated with a pronounced hypertensive phenotype in patients with adrenal CS that persisted even despite remission of hypercortisolism.

In subcutaneous adipose tissue the patients with CS disease did show more metabolic alterations in regard to lipogenesis and glucose oxidation [48], which was not commonly observed in the VAT in our study. However, we observed some similar effects including an increase in fatty acid synthase (FASN) and leptin (LEP) in CS patients. This could be due to the fact that the subcutaneous adipose tissue and visceral fat have strong differences in their metabolic activity, especially regarding lipolysis [49]. Our analysis indicated that adipocyte stem- and precursor cells were decreased in CS, as well as a strong regulation of important factors for adipocyte differentiation like adiponectin (ADIPOQ) and peroxisome proliferator-activated receptor-gamma (PPARG). This suggests an attenuated adipocyte differentiation and adipogenesis of precursor cells [50]. As patients with CS have a strong change in fat cell distribution and body weight gain [51], one of the reasons for this might be a changed adipose tissue identity caused by cortisol excess.

It has been proposed that in VAT of patients with CS, the hypercortisolism has strong effects on inflammatory genes and immune cell infiltration, with studies showing an increased number of infiltrating macrophages (CD68 positive cells) [52]. We could also observe and increased number of infiltrating monocytes and macrophages (**Figure 2A**). In contrast to other studies, we did not observe a low-grade inflammation but rather a suppression of inflammatory and increased expression of anti-inflammatory markers of macrophages and inflammation. The most common observation for inflammatory effects in the VAT of CS patients has been an increase in Cd11c (ITGAX) a marker for an increased proinflammatory state, which we did not observe in our patient cohort [41, 52]. In our study we did, however, see a strong downregulation of inflammatory cytokines like Il6 (IL6) and Cxcl3 (CXCL3) as well as a downregulation of the inflammatory NFKB pathway and MAPK pathways. This contrast could be explained that both studies showing an increase in ITGAX were sex mixed cohorts of patients, while our study only focuses on female patients.

A major challenge in patients with CS is the management of blood pressure [5, 53]. More so it was intriguing that we saw strong effects on endothelia cells and smooth muscle cell abundance, as well as important genes for the vascular system like Vegfa (VEGFA) and VWF (VWF). It has already been shown that cortisol excess can lead to an increase of thickness in arteries and therefore modulate systemic blood pressure and persistent modulation of the vascular system, even after achieving remission of endogenous cortisol excess [54-57]. While some of these vascular changes revert after the remission some persists, like the plaque formation [20, 58]. Furthermore, we saw a strong regulation of components of the RAAS in the VAT. For patients suffering from hypertension caused by obesity it was shown that local RAAS activity was a major factor in contribution to hypertension, irrespective of systemic regulation [59, 60]. In obesity, angiotensinogen levels are known to be elevated and VAT-derived angiotensinogen can enter the circulation, leading to an elevated blood pressure through the actions of angiotensin II. Here, the local activation of glucocorticoid action in adipose tissue of patients with obesity is proposed to induce activation of RAS [22]. Recently, a positive correlation between BMI and plasma angiotensinogen concentrations in patients with CS was observed [13].

We could further dissect this effect in correlating the VAT gene expression to patient features and show that within our group of CS patients the gene expression in VAT was correlated with the severity of the hypertensive phenotype (**Figure 3**). mRNA expression of Renin (REN) in VAT was strongly correlated with the systemic blood pressure (**Figure 4**). Interestingly this was independent of the severity of endogenous glucocorticoid excess.

The tissue RAS and the circulating RAS are in a state of constant interaction [22, 61]. Activation and/or modulation of the RAS is known to be a mechanism of glucocorticoid-induced hypertension in patients with CS [11, 16]. Despite elevated angiotensinogen (renin substrate) levels observed in CS [62], levels of renin activity, renin concentration, angiotensin II and aldosterone are reported in the normal or below the normal range [13, 43-46]. This is interpreted as an overall increased throughput of the RAS and/or the consequence of increased glucocorticoid-mediated mineralocorticoid receptor stimulation and thus renin suppression [13, 63]. Moreover, patients with CS showed an increased pressor response to angiotensin II [64, 65], and there is evidence that glucocorticoids increase angiotensin II type 1 receptor gene expression [66, 67]. Due to the observed generalized upregulation of the RAS in patients with CS, angiotensin-converting enzyme inhibitors or angiotensin receptor blockers are recommended as first-line approach for blood pressure control in CS [5].

In line with previous studies, our patients with active CS showed no significant difference in plasma renin and aldosterone concentrations compared to controls (**Figure 4C**). Interestingly, in the CS^High^ subgroup, we observed significantly decreased plasma renin concentrations compared to the the CS^Low^ subgroup. Since aldosterone and potassium levels are normal in both subgroups, showing no difference (**Figure 4C and S3D**), we assume a relative functional mineralocorticoid excess state in relation to blood pressure and systemic volume, which induces negative feedback regulation at kidney level decreasing systemic renin levels. This condition has also been described primarily in patients with ectopic CS [63, 68, 69]. Furthermore, several studies demonstrated that sodium reabsorption and blood volume expansion through mineralocorticoid receptor activation is insufficient to explain the development of glucocorticoid-induced hypertension [63, 70-73]. Since we observed a unique VAT gene expression profile with increased REN expression associated with a hypertensive phenotype and lower plasma renin levels in a subset of CS patients, we postulate a local overactivity of the RAS in VAT. This RAS activation, as suggested by others [13, 63], could lead to low plasma renin levels through increased angiotensin production and aldosterone synthesis, via a hypothesized negative feedback mechanism to limit excessive angiotensin II production [74].

Remarkably, in the study by van der Pas and colleagues [13], plasma concentrations of angiotensinogen, renin and aldosterone had not changed despite biochemical remission by medical therapy, indicating long-lasting effects on plasma RAS components that may be responsible for partial persistence of glucocorticoid-induced hypertension. Despite successful treatment of CS, about one third of patients have persistent hypertension [12, 20, 64]. As predictive factors for persistent hypertension, older age, higher BMI at baseline and a longer estimated duration of hypercortisolism were observed [17]. Of note, the severity of hypercortisolism does not seem to affect the severity of hypertension or the remission status of hypertension after successful treatment of CS [11-13, 17]. This is consistent with our observation that CS^High^ and CS^Low^ subgroup subgroups do not differ in the degree of hypercortisolism (**Figure 3B and S2B**). In our follow-up visits 6 months after adrenalectomy due to adrenal CS, a difference in blood pressure was still found between CS subgroups CS^High^ and CS^Low^, which may suggest an association of VAT RAS modulation and the persistence of glucocorticoid-induced hypertension. This could be either of epigenetic nature, as it has been previously reported that CS patients also show a changed epigenetic landscape or genetic nature exacerbated by the excess cortisol [41]. Here it was shown that markers for epigenetic gene activation (H3K27ac) and gene repression (H3K4me3) were altered during active CS.

We propose that local RAAS activity induced by local excess of cortisol, independent from systemic regulation could alter the systemic blood pressure and contribution to the hypertension observed in CS patients.

A limitation of our study is, that our patients used a variety of antihypertensive drugs, most of which are known to interfere with plasma concentrations of the RAS. However, plasma RAS components in patients with CS were not shown to be significantly affected by angiotensin-converting enzyme inhibitors, angiotensin receptor blockers or β-adrenoceptor blockers [13]. But of course, RAS blockade or β-adrenoceptor blockers could also have an effects of RAS components in VAT.

Why the subgroups of patients with CS determined on the basis of gene expression in VAT form remains unclear. Differences in local glucocorticoid levels, e.g., due to altered 11β hydroxysteroid dehydrogenase type 1 activity, are possible [63]. 11β hydroxysteroid dehydrogenase type 1 is found primarily in liver and adipose tissue and appears to have a role in arterial hypertension [63, 69, 75].

## Data Availability

Sequencing data was uploaded to the GEO database and is available under GSE218965. Other data will be available on request or deposited if possible.

## Author Contributions

US, JPT and MR conceptualized the study and planned experiments. US performed bioinformatic analyses. US and FV performed molecular biology assays. GC provided critical methods and protocols. JPT and MR supervised the work. US, FV, MR and JPT wrote the manuscript. All authors approved the manuscript.

## Acknowledgments

This work was funded by the Deutsche Forschungsgemeinschaft (DFG) within the collaborative Research Centre 1506 Aging at Interfaces at the Project C05 (*Project Number 450627322*) to J.T. M.R. received grant support for the German Cushing’s Registry CUSTODES by the Else Kröner-Fresenius Stiftung (2012_A103 and 2015_A228), by the Deutsche Forschungsgemeinschaft (DFG, German Research Foundation, Projektnummer: 314061271-TRR 205) and by the European Research Council under the European Union Horizon 2020 research and innovation program (grant agreement No. 694913). F.V. is supported by the Deutsche Forschungsgemeinschaft (DFG, German Research Foundation, project number: 413635475) and the Munich Clinician Scientist Program (MCSP) of the LMU München.

